# Application of Safety Attitudes Questionnaire (SAQ) in Adult Intensive Care Units: a cross-sectional study

**DOI:** 10.1101/2020.07.07.20114918

**Authors:** Abdullah S. Alqahtani, Rachel Evley

## Abstract

**Purpose:** To achieve a positive safety culture, staff perception of safety must be frequently measured. There are several active and reactive methods to use to measure safety cultures such as near-miss occurrence, accidental data collection, measuring behavior, self-report method, and safety questionnaires. The safety attitudes questionnaire (SAQ) tool was used to measure safety culture. This tool is widely used in literature and among researchers and has been used and validated in middle eastern cultures. In addition, it has a validated Arabic version.

**Methods:** A cross-sectional study was conducted using anonymous and random sampling. I surveyed all ICU staff working in all the adult ICUs in two of the major hospitals in the eastern province of Saudi Arabia. The short version of the Safety Attitudes Questionnaire was used to assess participants’ attitudes towards safety culture. The study involved all healthcare providers working in Adult ICU.

**Results:** The study occurred over a three-week period in March 2019. A total of 82 completed questionnaires were returned which represented a response rate of 82%. On average, the domain that scored the highest number of positive responses was Job satisfaction with 68.5%, followed by teamwork climate 67.8%, then working conditions 60.1%, 57.1% safety climate, then preparation of management with 53.4%, and finally 46% in Stress recognition. A statistically significant difference was found between the mean SAQ score and the educational level of the participants. Participants with bachelor’s degrees scored a mean of 50.17 compared to participants hold diploma degrees who scored a mean of 68.81 (*P*=0.02). Moreover, a significant difference was found between the mean SAQ score and participants’ specialties. Attending/Staff Physician mean score was 36.40, Nurse Manager/Charge Nurse scored 39.78, and Respiratory therapist mean score was 47.88, compared to mean score of 62.27 for Registered Nurse, and Respiratory supervisor 67.0 (*P*=0.04). In addition, 79.2% of the respondents did not report any incidents in the last 12 months.

**Conclusions:** The result of the study shows an unsatisfying level of safety culture among healthcare staff in ICUs. The importance of this study is to establish a baseline for safety climate in these hospitals and specifically ICUs. In addition, by exposing the system weaknesses it helps the administration to strengthen and improve patient care. By decreasing workload and job stress, studies show they have a positive association with increasing job performance.

## Introduction

Patient safety, prevention of medical error, and quality of care are important aspects of health care. The importance of these fields has led to an expansion in literature and research in recent years. As in any other profession or industry such as engineering, aviation, or nuclear plants, human factors play an important role in the generation of medical incidents (Reason 2000). The available literature seems to suggest that high a percentage of medical incidents could be prevented in many cases (Brennan et al. 1991).

To achieve a positive safety culture, first, was must understand the values, perceptions, and patterns of behaviors to determine what is the norms and what is appropriate. After the Chernobyl disaster in 1986, more attention was given to safety culture in nuclear institutions. In 1993, the Advisory Committee on the Safety of Nuclear Installations (ACSNI) established a definition for safety culture as: “*The safety culture of an organization is the product of individual and group values, attitudes, perceptions, competencies, and patterns of behavior that determine the commitment to, and the style and proficiency of, an organization’s health and safety management*.

*Organizations with a positive safety culture are characterized by communications founded on mutual trust, by shared perceptions of the importance of safety and by confidence in the efficacy of preventive measures*.*”* (Health and Safety Commission (HSC) 1993).

Safety culture in a unit is a part of the whole organizational culture. Cooper described safety culture as a sub-facet of a bigger culture, the organizational culture (Cooper 2000). Moreover, cooper also suggests that safety culture affects organization members’ attitudes and behavior about the organization’s current health and safety performance (Cooper 2000). As well, the administration’s perception of safety influences the staff safety culture (Gadd and Collins 2002). Communication-based on mutual trust and committing and understanding the importance and benefits of following safety protocols are features of positive safety culture (Sorra and Nieva 2004).

### In the Cooper framework, safety culture has three aspects

Psychological aspect, Behavioral aspect, and Situational aspect. The psychological aspect (This is also called the safety climate) which represents the individual’s attitude, values, and perceptions. It can be measured by the safety climate questionnaire to capture a glimpse of staff attitude and perceptions toward safety at the point of time. However, in some studies, safety climate has come to be used to refer to safety culture alternatively (Gabrani et al. 2015; Soh et al. 2016). In some studies, safety climate has been described as the shared perceptions, attitudes, and beliefs of staff about the organization handle and archives safety (Flin et al. 2006; Soh et al. 2016). To reiterate Safety culture is how things are done around here, and safety climate is a glimpse or superficial image of safety culture in a specific time and place. However, throughout this paper, the term safety climate will be used to refer to the psychological aspect of safety culture.

The second aspect: the behavioral aspect, which focuses on individuals’ actions can be examined by direct peer observations and self-reports.

Third aspect: the situational aspect, concerns the organization’s policies, procedures, regulations, and organizational system. This aspect could be examined via inspections and surveillance (Cooper 2000; Health & Safety Executive 2005).

As in different industries such as aviation and nuclear power, the Agency of Health Care Research and Quality (AHRQ) (Sorra et al. 2016), the Joint Commission for the Accreditation of Healthcare Organizations (Joint Commission 2018), and the National Health Service (NHS) (NHS England and NHS Improvement 2019) encourage the regulatory measurement of safety culture. To maintain or to improve safety culture it must be frequently measured. Measuring safety could be done actively or reactively.

Active measurement is done to monitor safety culture in the organization before an incident occurs; these measurements are done as inspections rather than investigation. On the other hand, reactive measurement is measuring safety culture after an incident took place, investigating the incident or near miss cases, and examining the safety culture at that moment (Gadd and Collins 2002).

There are several active and reactive methods to use to measure safety cultures such as near-miss occurrence, accidental data collection, measuring behavior, self-report method, and safety questionnaires.

One of the common methods to actively assess safety culture is safety questionnaires and one of the most internationally used questionnaires is the Safety Attitude Questionnaire (SAQ) (Gabrani et al. 2015; Smits et al. 2017). SAQ was developed by the University of Texas and partially funded by AHRC, with the aim to fill the demand for healthcare quality regulations from the Joint Commission for the Accreditation of Healthcare Organizations, the Agency for Health-care Research and Quality, and the U.S. National Quality to measure and monitor safety culture (Sexton et al. 2006).

SAQ has many strengths, it is popular among researchers and investigators, has a shorter questionnaire compared to others such as The Job Descriptive Index, tested and validated in different cultures worldwide, available in multi-language translations, ability to measure and monitor trend data over time, and ability to benchmark and utilize it within any unit in the hospital such as ICU and OR. (Etchegaray and Thomas 2012; Gabrani et al. 2015; Smits et al. 2017) SAQ ICU Arabic version is the most used validated tool in Arabic culture. SAQ was translated and validated (internal consistency) by Hamdan 2013 (Hamdan 2013). Moreover, Abu-El-Noor et al in the 2017 study confirmed Hamdan translation’s, tested the psychometric properties (validity and reliability) and found good validation, they determined that it could be used as a tool to evaluate the safety attitudes in Arabic speaking hospital culture (Abu-El-Noor et al. 2017).

## Method

A cross-sectional study was conducted using anonymous and random sampling. I surveyed all ICU staff working in all the adult ICUs in two of the major hospitals in the eastern province of Saudi Arabia. The short version of the Safety Attitudes Questionnaire was used to assess participants’ attitudes towards safety culture. Approval for the Questionnaire was gained from the University of Texas for the English version. Permission was obtained from Dr. Hamdan for the validated Arabic version of the Questionnaire.

It has 30 core items representing six scales: teamwork climate, safety climate, job satisfaction, stress recognition, perception of management, and working conditions. Additional items were added to explore the safety culture perception and the attitude toward incident reporting within the unit.

The following items were not part of the original Questionnaire: Item “How you score the intensive care unit in this hospital regarding patient safety”, item “describe the quality of communication and collaboration with the following personnel during your experience in this ICU”, and item “How many incidents (medical errors) did you report to the management in the last 12 months?”.

The questionnaire uses a Likert-type scale, ranging from 5 for strongly agree, 4 slightly agree, 3 neutral, 2 slightly disagree, to 1 for strongly disagree. The overall domain score was calculated by summing all items in each domain then divided by the number of items.

### Data collection

Data collection for the study occurred over a three-week period in March 2019. The respondent criteria included all healthcare workers in the adult intensive care units with at least one year of experience and worked in the ICU for at least one month before the study.

### Statistical Analysis

All data were transferred to an electronic worksheet in Microsoft® Excel on office 365. Then data were analyzed by using SPSS software (version 25, Chicago, IL). A p-value of less than 5% was considered to be statistically significant. Cronbach’s alpha was used to assess the reliability of each domain in the Questionnaire. Descriptive analysis was performed using ANOVA and paired sample t-tests in all demographic data including sex, age, educational level, job title, years of experience, working hours and working shifts and compared to SAQ mean scores.

## Results

A total of 82 completed questionnaires were returned which represented a response rate of 82%. **Table 1** shows the demographics of the questionnaire participants. 49% of the participants used the Arabic version of the Questionnaire versus 51% using the English version. 57% of the participants were male, and 43% were female. Among participants, around 45% were respiratory therapists, 45% were nurses, 9% were physicians, and one participant was an ICU nutrition specialist. Most of the participants were bachelor’s degree holders (76%), then diploma (18%), and postgraduates (6%). 63% of participants have a minimum of 5 years of experience. Regarding working hours, the majority were working 40 to 59 hours per week (83%), and the part-time staff was 11.7%. Around 72% of the participants’ work schedule is variable and they cover night and day shifts, 22% work only day shifts, and 6% work night shifts.

**Table 1:** Demographic data of respondents.

| **Variables** | **Frequency** | **Percentage** |
| --- | --- | --- |
| **Sex (n= 79)** |  |  |
| Male | 45 | 57 |
| Female | 34 | 43 |
| **Age (n = 75)** |  |  |
| <= 30 Years | 40 | 53.3 |
| > 30 Years | 35 | 46.7 |
| **Educational Level (n=79)** |  |  |
| Diploma | 14 | 17.7 |
| Bachelor's degree | 60 | 75.9 |
| Master, Ph.D., or equivalent | 5 | 6.3 |
| **Job Title (n=78)** |  |  |
| Staff Physician | 5 | 6.4 |
| Nurse | 32 | 41.0 |
| Resident Physician | 1 | 1.3 |
| Respiratory Therapist | 33 | 42.3 |
| ICU Head | 1 | 1.3 |
| Nurse In-charge | 3 | 3.8 |
| Respiratory Supervisor | 2 | 2.6 |
| Nutrition Specialist | 1 | 1.3 |
| **Years in Profession (n= 79)** |  |  |
| < = 5 Years | 29 | 36.7 |
| > 5 Years | 50 | 63.3 |
| **Years in ICU (n= 79)** |  |  |
| < = 5 Years | 43 | 54.4 |
| > 5 Years | 36 | 45.6 |
| **Work hours per week (n = 77)** |  |  |
| Less than 39(part-time) | 9 | 11.7 |
| 40 to 59 hours | 64 | 83.1 |
| 60 to 79 hours | 2 | 2.6 |
| 80 hours and more | 2 | 2.6 |
| **Work shift (n = 79)** |  |  |
| Day | 17 | 21.5 |
| Night | 5 | 6.3 |
| Variable shifts | 57 | 72.2 |
| **Number of incidents reported to the Management (n = 80)** |  |  |
| None | 62 | 77.5 |
| 1 – 2 incidents | 18 | 22.5 |
| **Language of survey (n=82)** |  |  |
| Arabic | 40 | 48.8 |
| English | 42 | 51.2 |

The participant’s mean scores for each item of the six domains and the percentage of positive responses are presented in **Table 2**. On average, the domain that scored the highest number of positive responses was Job satisfaction with 68.5%, followed by teamwork climate 67.8%, then working conditions 60.1%, 57.1% safety climate, then preparation of management with 53.4%, and finally 46% in Stress recognition. **Figure 1** demonstrates the average percentage of positive responses per SAQ domain.

**Table 2:** SAQ Domains and items, scales’ mean scores, and percentages of positive responses.

|  | Mean | SD | % of Positive Responses |
| --- | --- | --- | --- |
| **Job satisfaction (Cronbach's $\alpha = 0.86$)** |  |  |  |
| I like my Job | 3.56 | 2.07 | 75.7 |
| Working here is like being part of a large family | 3.07 | 2.20 | 67.0 |
| This is a good place to work. | 3.04 | 2.24 | 65.8 |
| I am proud to work in this clinical area. | 3.38 | 2.10 | 73.1 |
| Morale in this clinical area is high. | 2.72 | 2.22 | 60.9 |
| **Safety Climate (Cronbach's $\alpha = 0.77$)** |  |  |  |
| I would feel safe being treated here as a patient | 2.40 | 2.33 | 52.5 |
| Medical errors are handled appropriately in this clinical area | 2.38 | 2.36 | 51.2 |
| I know the proper channels to direct questions regarding patient safety in this clinical area. | 3.02 | 2.17 | 67.0 |
| I receive appropriate feedback about my performance. | 2.99 | 2.09 | 68.3 |
| In this clinical area, it is difficult to discuss errors. | 1.48 | 2.02 | 35.4 |
| I am encouraged by my colleagues to report any patient safety concerns I may have. | 2.83 | 2.14 | 64.6 |
| The culture in this clinical area makes it easy to learn from the errors of others. | 2.68 | 2.19 | 61.0 |
| **Teamwork climate (Cronbach's $\alpha = 0.68$)** |  |  |  |
| Nurse input is well received in this clinical area | 3.37 | 2.09 | 73.2 |
| In this clinical area, it is difficult to speak up if I perceive a problem with patient care | 1.00 | 1.85 | 23.2 |
| Disagreements in this clinical area are resolved appropriately (i.e., not who is right, but what is best for the patient). | 2.65 | 2.27 | 58.5 |
| I have the support I need from other personnel to care for patients | 3.43 | 2.07 | 74.4 |
| It is easy for personnel here to ask questions when there is something that they do not understand. | 3.45 | 2.02 | 75.6 |
| The physicians and nurses here work together as a well-coordinated team. | 3.39 | 2.11 | 73.1 |
| **Working conditions (Cronbach's $\alpha = 0.84$)** |  |  |  |
| Problem personnel are dealt with constructively by our department. | 2.09 | 2.23 | 47.6 |
| This hospital does a good job of training new personnel. | 2.82 | 2.25 | 62.2 |
| All the necessary information for diagnostic and therapeutic decisions is routinely available to me. | 2.93 | 2.16 | 65.9 |
| Trainees in my discipline are adequately supervised. | 2.95 | 2.23 | 64.6 |
| **Preparation of Management (Cronbach's $\alpha = 0.70$)** |  |  |  |
| Management supports my daily efforts: | 2.24 | 2.18 | 52.4 |
| Management doesn't knowingly compromise patient safety. | 2.76 | 2.31 | 59.8 |
| I get adequate, timely info about events that might affect my work. | 2.44 | 2.20 | 56.1 |
| The levels of staffing in this clinical area are sufficient to handle the number of patients | 1.98 | 2.22 | 45.1 |
| **Stress recognition (Cronbach's $\alpha = 0.71$)** |  |  |  |
| When my workload becomes excessive, my performance is impaired. | 2.05 | 2.24 | 46.3 |
| I am less effective at work when fatigued. | 2.54 | 2.18 | 58.5 |
| I am more likely to make errors in tense or hostile situations. | 1.71 | 2.23 | 37.8 |
| Fatigue impairs my performance during emergency situations (e.g. emergency resuscitation, seizure). | 1.85 | 2.24 | 41.5 |
| Overall SAQ (Cronbach's $\alpha = 0.91$) | 2.64 | 1.13 | |

**Figure 1:**
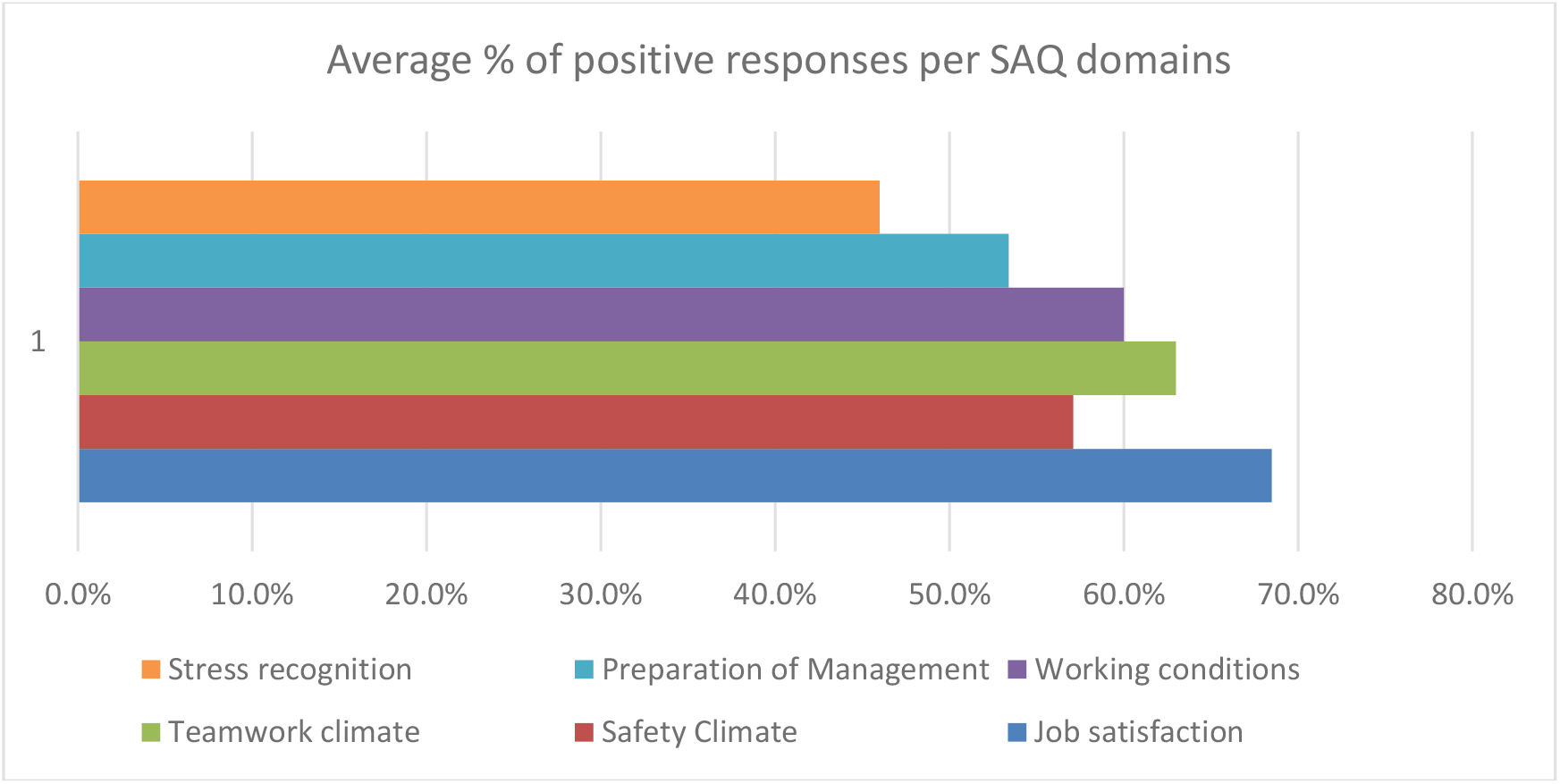
Average of percentage positive responses per SAQ domains.

The Pearson correlation analysis was used to determine the associations between participants’ positive response and each SAQ domain, the reliability analysis was done using Cronbach α method and showed that the Cronbach α values.

Overall, the SAQ had very good internal consistency (Cronbach’s α = 0.91). The domains internal consistency showed good results for Job satisfaction (Cronbach’s α = 0.86), Safety Climate (Cronbach’s α = 0.77), Working conditions (Cronbach’s α = 0.84), and Stress recognition (Cronbach’s α = 0.71). And acceptable internal consistency in Preparation of Management (Cronbach’s α = 0.70), and Teamwork climate (Cronbach’s α = 0.68) which fall within the acceptable range (De Vet et al. 2011).

**Table 3** shows the mean SAQ score of participants according to their characteristics. A statistically significant difference was found between the mean SAQ score and the educational level of the participants. Participants with bachelor’s degrees scored a mean of 50.17 compared to participants hold diploma degrees who scored a mean of 68.81 (*P*=0.02). Moreover, a significant difference was found between the mean SAQ score and participants’ specialties. Attending/Staff Physician mean score was 36.40, Nurse Manager/Charge Nurse scored 39.78, and Respiratory therapist mean score was 47.88, compared to mean score of 62.27 for Registered Nurse, and Respiratory supervisor 67.0 (*P*=0.04).

**Table 4** shows the mean SAQ score of both hospitals in each domain. Perception of management scored a mean of 28.65 in hospital 2 compared to 62.22 in hospital 1. Hospital 2 working condition scored a mean of 29.05, compared to 74.33 in hospital 1. Moreover, the safety climate mean score in hospital 2 was 37.22, and hospital 1 was 69.97. This could be explained by the significant difference in management and working conditions in both hospitals. Hospital 1 is a military hospital that is managed by the Ministry of Defense and serves only the Ministry of Defense workers and their families. Whereas Hospital 2 is under the Ministry of Health administration and serves the public residents of the region and is therefore expected to have a higher workload compared to hospital 1. In addition, even though the bed capacity of hospital 2 (425 beds) is more than hospital 1 (335 beds) the staffing in hospital 2 is insufficient and therefore working conditions and perception of management mean score was immensely negative.

One of the added items to the Questionnaire was the number of incidents reported by the participants in the last 12 months. Based on the responses, **table 5** shows 79.2% of the respondents did not report any incidents in the last 12 months. 20.7% report one or two incidents in the last 12 months and no participant have reported more than two incidents (**figure 2**).

**Table 3:** Association between participant characteristics and overall SAQ scores.

|  | Mean | SD | P-value |
| --- | --- | --- | --- |
| Sex (n= 79) |  |  | 0.43 |
| Male | 55.08 | 23.23 |  |
| Female | 52.10 | 20.70 |  |
| Age (n = 75) |  |  | 0.30 |
| <= 30 Years | 51.22 | 20.94 |  |
| > 30 Years | 56.42 | 23.49 |  |
| Educational Level (n=79) |  |  | 0.02 |
| Diploma | 68.81 | 17.26 |  |
| bachelor's degree | 50.17 | 22.17 |  |
| Master, Ph.D., or equivalent | 55.33 | 17.48 |  |
| Job Title (n=78) |  |  | 0.04 |
| Attending/Staff Physician | 36.40 | 8.39 |  |
| Registered Nurse | 62.27 | 22.30 |  |
| Respiratory therapist | 47.88 | 21.09 |  |
| Fellow Physician | 70.00 | nil |  |
| Nurse Manager/Charge Nurse | 39.78 | 13.68 |  |
| Unit Head | 67.33 | nil |  |
| Respiratory supervisor | 67 | 0.47 |  |
| Nutrition | 78.67 | nil |  |
| Years in Profession (n= 79) |  |  | 0.65 |
| < = 5 Years | 55.29 | 21.62 |  |
| > 5 Years | 52.93 | 22.52 |  |
| Years in ICU (n= 79) |  |  | 0.24 |
| < = 5 Years | 56.50 | 23.10 |  |
| > 5 Years | 50.57 | 20.66 |  |
| Work hours per week (n = 77) |  |  | 0.94 |
| Less than 39(part-time) | 53.85 | 26.24 |  |
| 40 to 59 hours | 53.42 | 21.98 |  |
| 60 to 79 hours | 43.33 | 5.66 |  |
| 80 hours and more | 52.00 | 25.46 |  |
| Work shift (n = 79) |  |  | 0.28 |
| Day | 55.61 | 20.42 |  |
| Night | 38.53 | 18.05 |  |
| Variable shifts | 54.60 | 22.68 |  |
| Number of incidents reported to the Management (n = 80) |  |  | 0.21 |
| None | 52.12 |  |  |
| 1 – 2 incidents | 59.48 |  |  |

**Table 4:** Comparison between the result of both hospitals.

| Domain | Hospital 1 |  | Hospital 2 |  |
| --- | --- | --- | --- | --- |
|  | Mean | SD | Mean | SD |
| Job Satisfaction | 76.71 | 30.17 | 46.49 | 32.41 |
| Safety Climate | 61.97 | 25.21 | 37.22 | 25.93 |
| Teamwork Climate | 66.67 | 21.09 | 46.58 | 26.88 |
| Working Conditions | 74.33 | 27.11 | 29.05 | 30.66 |
| Perception of management | 62.22 | 25.73 | 28.65 | 29.12 |
| Stress recognition | 35.44 | 28.16 | 47.16 | 36.22 |

**Table 5:** Number of incidents reported by participants per specialty in the last 12 months.

| # of incidents reported | All participants |  | Physicians |  | Nurses |  | RT |  | Others |  |
| --- | --- | --- | --- | --- | --- | --- | --- | --- | --- | --- |
|  | F | % | F | % | F | % | F | % | F | % |
| 1 – 2 | 17 | 20.7 | 3 | 3.7 | 7 | 8.5 | 6 | 7.3 | 1 | 1.2 |
| None | 65 | 79.2 | 3 | 3.7 | 28 | 34.1 | 29 | 35.4 | 5 | 6.1 |

**Figure 2:**
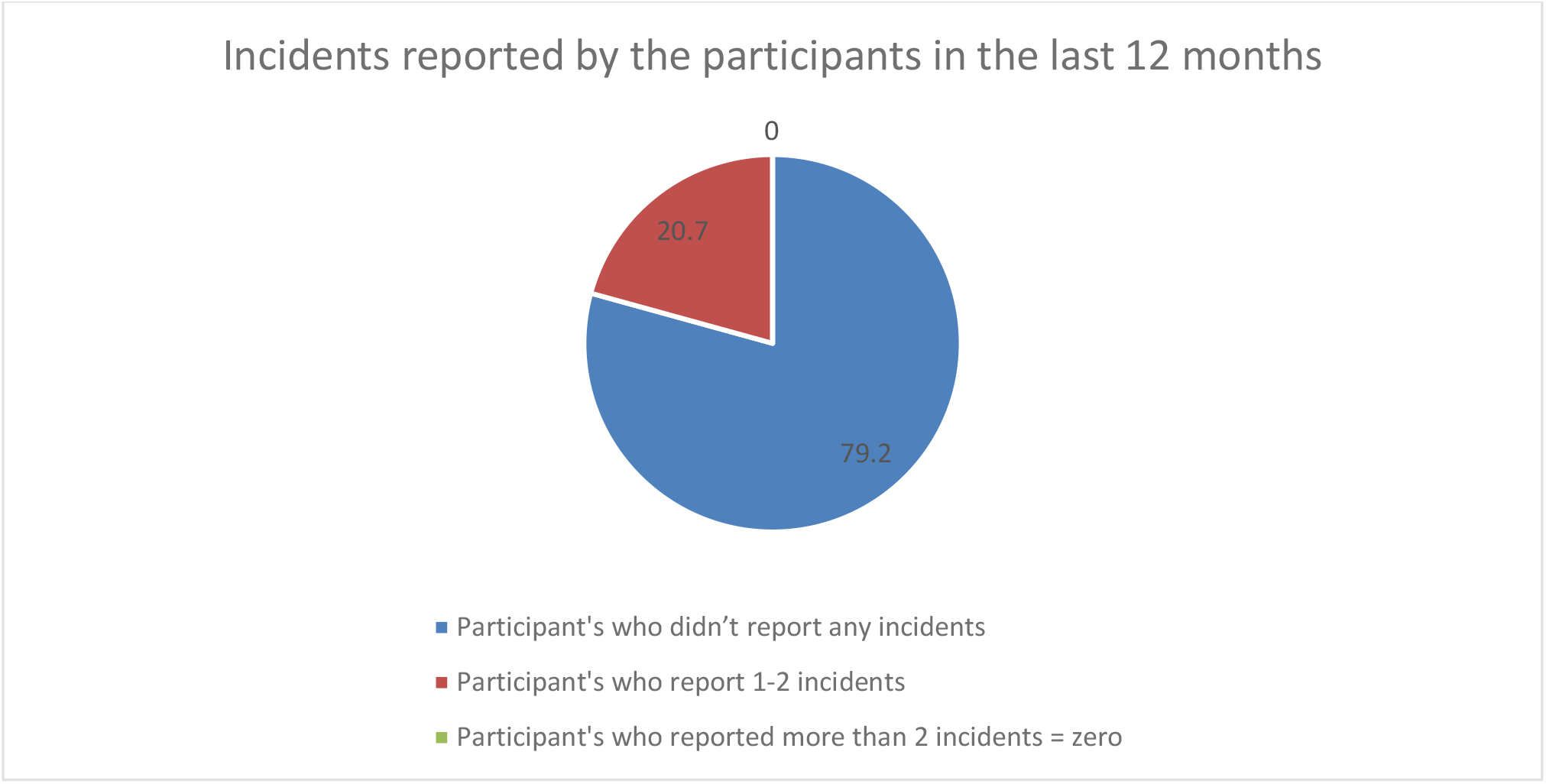
Number of incidents reported by participants in the last 12 months.

## Discussion

Healthcare accreditation organizations such as Joint Commission International (JCI), United Kingdom Accreditation Service (UKAS) and the Saudi Central Board for Accreditation of Healthcare Institutions (CBAHI), encourage hospitals to frequently assess safety culture within the institution. In Saudi Arabia, the safety culture research field is still relatively new and under-explored and has very limited published data in measuring safety culture and the use of SAQ (Alayed, Lööf, and Johansson 2014; Algahtani 2015; Alswat et al. 2017; Alzahrani, Jones, and Abdel-Latif 2018). The findings of this study show similarity to results from other studies (Abu-El-Noor et al. 2017; Gabrani et al. 2015; Hamdan 2013; Sexton et al. 2006; Vifladt et al. 2016; Zhao et al. 2019).

In this study, results show a significant difference between nurses and other specialties. Nurses scored a higher positive attitude toward safety culture. The result is in line with the findings from the Abu-El-Noo study (Abu-El-Noor et al. 2017). This difference could be related to the fact that most of the nurses were younger and felt more informed toward safety culture, they may also be more resilient and able to cope with stressful working conditions in ICU (Raftopoulos and Pavlakis 2013a). However, other studies reported that physicians scored higher than nurses (Gabrani et al. 2015; Sexton et al. 2006; Thomas, Sexton, and Helmreich 2003). The difference in SAQ scores between healthcare specialties could be related to safety culture inside their department, responsibilities, availability of protocols, training, and gender (Thomas et al. 2003).

Another significant difference was found between staff holding diploma degree who scored higher compared to higher educated staff. This could be related to the fact that most of the diploma staff were working in their ICUs for a longer time than others and they were more familiar with the unit policies and team. Findings in this study indicate that staff working in night shifts scored lower overall SAQ than the day shift. Various studies found that safety and productivity are compromised during night shifts (Folkard and Tucker 2003; Gomez-Garcia et al. 2016; Wagstaff and Lie 2011).

In this study, job satisfaction was scored the highest positive domain among all domains despite the fact that the stress recognition domain scored the lowest 46% and more than 55% of participants believe the ICU staffing is insufficient. However, these results were consistent with findings from other studies (Hamdan 2013; Raftopoulos and Pavlakis 2013b). This could be explained by firstly, the high working morale since 75.7% of participants answered positively to the item “I like my Job” which was the highest scored item in SAQ. Secondly, by the positive teamwork domain, which scored 67.8% positive response (2^nd^ highest domain) which could be a result of ICU staff familiarity with each other. Moreover, it was noticed that some of the young staff were trained during their internship in these units; therefore, they feel loyal to the team.

The stress recognition domain scored the lowest in both hospitals. This finding is consistent with other studies (Raftopoulos and Pavlakis 2013b). This could be a result of working in a stressful, fast-paced environment, and staff been burned out. In addition, Item “Problem personnel are dealt with constructively by our department” in the working condition received only 47.6% positive response. Several studies showed a correlation between high workload, stressful working conditions, and the number of incidents rate (Abu-El-Noor et al. 2017; Ahola et al. 2009; Fahrenkopf et al. 2008; Guirardello 2017).

The results of overall SAQ show a significant deferent between both ICUs. However, the variation in overall SAQ score between hospitals is common and expected as a result of different management, financial status, staff level, and type of patients (Hamdan 2013; Raftopoulos and Pavlakis 2013b).

Hospital 1 had recently initiated a safety educational program among the hospital staff, which could explain the relatively positive attitude of their staff toward safety culture. However, both hospitals showed a lack of proper reporting system and staff unwilling to report incidents (**Table 5**). This could be a result of many reasons such as lack of staff awareness about the importance of reporting incidents, or fear of punishment and liability. In the survey 35.4% of participants had felt that: “In this clinical area, it is difficult to discuss errors” and 23.2% of participants felt that: “In this clinical area, it is difficult to speak up if I perceive a problem with patient care”, moreover only 51.2% of participants believes that “Medical errors are handled appropriately in this clinical area”. which indicates that the existing culture in the ICUs is not supportive and reliable toward reporting incidents and patient safety culture. Only 52.5% of staff said they felt safe to be treated in their hospitals, which is a strong indicator of poor patient safety.

Several studies showed that safety culture has a strong association with patient safety and incidents rate (Lee et al. 2010, 2019; Nieva and Sorra 2003; Pettker et al. 2011; Sexton et al. 2006; Tear et al. 2020). Moreover, more studies suggest that inability or difficulty for staff to report or to discuss incidents is one of the main reasons for poor patient safety culture (Buljac-Samardzic, Van Wijngaarden, and Dekker-Van Doorn 2016; Sexton et al. 2006). Therefore, further investigation is needed from both hospital administration to highlight the staff concerns and work in improving the safety culture.

## Conclusion

The result of the study shows an unsatisfying level of safety culture among healthcare staff in these 2 ICUs. The importance of this study is to establish a baseline for safety climate in these hospitals and specifically ICUs. In addition, by exposing the system weaknesses it helps the administration to strengthen and improve patient care. By Decreasing workload and job stress, studies show they have a positive association with increasing job performance (Raziq and Maulabakhsh 2015). The use of questionnaires should be done periodically to assess safety culture over a period of time and observe the direction of the results.

## Limitations

Using a questionnaire to evaluate safety culture or to be specific safety climate, plays an important role in drawing the road map of the institution safety culture assessment. However, SAQ is not enough, other tools should be used to inspect other aspects of safety culture. SAQ measures staff beliefs about safety culture rather than their actual safety behavior (Gadd and Collins 2002). Furthermore, the sample size was limited; therefore, the findings should be dealt with caution in regard to transferability. In addition, the participation of specialties such as physicians, clinical pharmacists, and others was poor and needed to be further investigated.

## Data Availability

The datasets generated during and/or analyzed during the current study are available from the corresponding author on reasonable request.

